# Endothelial receptor proteins in acute venous thrombosis and delayed thrombus resolution in cerebral sinus vein thrombosis

**DOI:** 10.1101/2023.11.06.23297882

**Authors:** Lukas Kellermair, Christoph Höfer, Matthias W.G. Zeller, Christa Kubasta, Dave Bandke, Serge Weis, Jörg Kellermair, Thomas Forstner, Raimund Helbok, Milan R. Vosko

## Abstract

**Background and Purpose:** Cerebral sinus venous thrombosis (CSVT) is a rare but life-threatening disease and its diagnosis remains challenging. Blood biomarkers, including D-Dimer are currently not recommended in guidelines. Soluble endothelial receptor proteins (sICAM-1, sPECAM-1 and sVCAM-1) have been shown to be promising diagnostic biomarkers in deep-vein-thrombosis (DVT) and pulmonary embolism (PE), however, their role in acute CSVT remains unclear.

**Methods:** In this bi-center, prospective study we quantified D-Dimer as well as sICAM-1, sPECAM-1 and sVCAM-1 in plasma of patients with clinically suspected CSVT managed in the neurological emergency department (ED) of a tertiary care hospital. All patients underwent cerebral magnetic resonance imaging (MRI) and were followed up after 3, 6 and 12 months to detect thrombus resolution.

**Results:** Twenty-four out of 75 (32%) patients with clinically suspected CSVT presenting with headache to the ED were diagnosed with acute CSVT. These patients had a mean age of 45 ± 16 years and 78% were female. In patients with CSVT, mean baseline D-dimer (p<0.001) and sPECAM-1 (p<0.001) were significantly higher compared to patients without CSVT. The combination of D-Dimer and sPECAM-1 yielded the best ROC-AUC (0.994; □ < □ 0.001) with a negative predictive value of 95.7% and a positive predictive value of 95.5%. In addition, higher baseline sPECAM-1 levels (> 198ng/ml) on admission were associated with delayed venous thrombus resolution at 3 months (AUC = 0.83).

**Conclusion:** sPECAM-1 in combination with D-Dimer should be used to improve the diagnostic accuracy of acute CSVT and sPECAM-1 may predict long-term outcome of CSVT. Confirmatory results are needed in other settings in order to show their value in the management concept of CSVT patients.

## Introduction

Cerebral sinus venous thrombosis (CSVT) is a rare form of venous thromboembolism (VTE) and is challenging to diagnose due to its highly variable clinical manifestations and the absence of reliable biomarkers. Recent studies have shown that the incidence of CSVT is higher than expected ^1,2^, which may be the result of the use and availability of more advanced diagnostic techniques. CSVT frequently occurs in young people, women of childbearing age, and children. ^3-5^ The clinical features of CSVT are variable and unspecific. Up to 80% of CSVT cases have an acute to subacute onset.^4^ The leading presenting clinical symptom is headache which can range from thunderclap-like types to migrainous and tension like headaches.^6^ Focal neurological symptoms such as visual loss and sensorymotor deficits are common. In 25% of patients, an isolated headache is the only symptom. Although prognosis is good in most patients, a significant number (about 13%) suffers from residual disabilities or death. ^6,7^

A frequently-discussed biomarker for predicting CSVT is D-Dimer. D-Dimer results from fibrin degradation via fibrinolysis and can predict VTE.^8^ However, false □ negative D □ dimer are frequent and lead to unnecessary imaging. ^9-12^

PECAM-1, also denoted CD 31, is a cell surface receptor-protein expressed on endothelial cells which has important roles in inflammation, angiogenesis, and thrombus resolution. ^13,14^ Intracellular adhesion molecule 1 (ICAM-1), also named CD 54, is another adhesion molecule which is involved in thrombosis and the development of post-thrombotic-syndromes (PTS). ^15,16^ Vascular cell adhesion molecule 1 (VCAM-1) is essential for leukocyte recruitment and the development of arteriosclerosis. Its role in venous thrombosis is unknown.^17,18^. Soluble endothelial receptor proteins have been shown to be promising biomarkers for deep vein thrombosis (DVT) and pulmonary embolism (PE).^13,19^ In this study we aimed to investigate the role of endothelial receptor proteins (PECAM-1; ICAM-1 and VCAM-1) in acute CVST and hypothesized that each biomarker, or a combination of these biomarkers can be used to predict acute CSVT and could be higher in CVST patients with delayed thrombus resolution.

## Methods

### Study Population

The study was approved by the local ethical review board (Ethik-Kommission Land Oberösterreich; EK-58-17). Patients with clinically suspected CSVT were screened and enrolled in the outpatient department for neurological emergencies of the Kepler University Hospital and of The Hospital of the Brothers of Saint John of God in Linz, Austria. All patients were referred to one of the hospitals with newly acute (1-2 days) or subacute (up to 10 days) headache, with or without focal neurological disabilities or seizures, and clinically suspected CSVT (headache and additional risk factors for VTE). All patients underwent a detailed neurological examination. Medical history, current medication and risk factors for VTE (previous deep venous thrombosis [DVT] immobilization, surgery, malignancy, pregnancy, hormone therapy, nicotine, body mass index (BMI), thrombophilia, recent travel, age, and gender) were recorded prospectively in all patients. Recruitment process is presented inFig.1. Patients without CSVT and clinical stable conditions were released from the hospital. Patients with CSVT were hospitalized.

**Fig.1.**
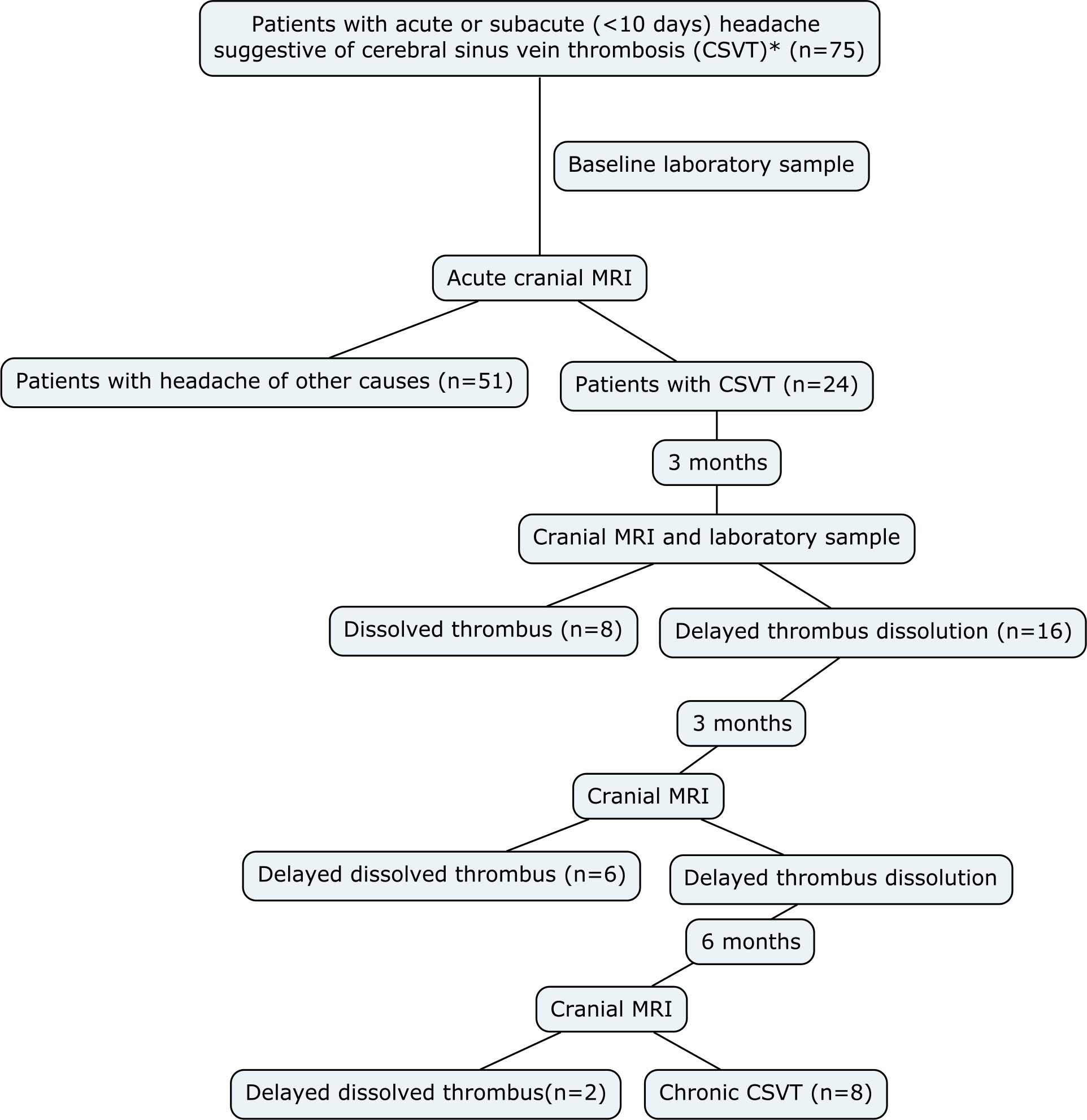
Flowchart of participants *Suggestive for cerebral sinus vein thrombosis were patients with newly or subacute headache with conspicuous anamnesis, neurological deficits or risk factors for vein thrombosis (summarized in Table.1)

Chronic headache was defined as headaches for at least 3 months based on ICD-10 criteria.^20^

### Blood samples

All patients underwent routine blood laboratory analysis including D-Dimer (mg/L) and blood samples for sPECAM-1 (ng/mL), sICAM-1(ng/mL) and sVCAM-1 (ng/mL) at baseline, and, if CSVT was diagnosed, additionally after 3 months (± 4 weeks). Blood samples for quantification of sPECAM-1, sICAM-1 and sVCAM-1 were immediately centrifuged at 4 °C, 3000×g for 10 min and stored at □ − □ 80 °C until final analysis. Measurements were then performed using ELISA (sandwich platinum instant ELISA-Kit; Thermo Fisher science, San Diego, USA) according to manufacturer’s instructions.

A D-Dimer cut-off value of >0.5 mg/L was defined as abnormal.^11,21,22^ ΔD-Dimer, ΔsPECAM-1, ΔsICAM-1 and ΔsVCAM-1 were defined as the difference between baseline and 3 months folllow-up (FU1).

### Neuroimaging

All patients underwent cranial MRI (1.5 Tesla, SIEMENS MAGNETOM Avanto syngo MR B19) at baseline. An additional scan was done after 3 months in patients with CSVT. All patients with delayed thrombus resolution underwent cranial MRI every 3 months (± four weeks) for up to 12 months. Delayed thrombus resolution was defined when residual thrombus was still visible after 3 months brain MRI despite anticoagulant treatment. “Chronic sinus thrombosis” was defined as still significant thrombotic residues after 12 months in neuroimaging.

The standardized protocol included an axial T2w-FLAIR sequence and a high-resolution T1-weighted 3D-MPRAGE sequence as well as T2-weighted, diffusion tensor images, susceptibility weighted imaging and venography.

### Treatment

All patients with CSVT were admitted to a stroke unit or neurological intensive care unit and were managed following international guidelines. The initial treatment included adjusted-dose of unfractionated heparin (UFH) or weight-based low-molecular-weight-heparin (LMWH; 1mg/kg twice daily) followed by vitamin K antagonists (or Off-Label Dabigatran 150 mg twice a day), regardless of the presence of intracerebral hemorrhage. One pregnant patient received therapeutic LMWH for 6 months and subsequent prophylactic LMWH. All patients with acute symptomatic seizures were treated with antiepileptic drugs.^6,23^

## Statistics

Sample size calculation was based on preliminary data^13^ using the sample size calculator provided by the Department of Biometrics, University of Münster, Germany for the primary endpoint sPECAM-1 using a power of 90% at a type I error of 5%.

Continuous data are presented as mean and standard deviation, categorical data are presented using counts and percentages. Normally distributed continuous variables were compared using the Student’s t-test or the Welch’s t-test in case of variance heterogeneity (verification with Levene’s test). The exact Mann-Whitney-U test was applied in case of non-normally distributed continuous variables. Assumptions of normal distribution for continuous variables were tested with the Kolmogorov-Smirnov test with Lilliefors correction. For categorical data the exact Chi-square test (n x k tables) or Fisher’s exact test (2 x 2 tables) were used.

Binary logistic regression was used to analyze variables (D-Dimer/sPECAM-1/sICAM-1/sVCAM-1/the combination of these variables) with regard to CSVT status. For various cut-off points distinct sensitivity/specificity values were calculated, receiver operating characteristic curves (ROC) were constructed, and ROC-AUC (area under curve) including 95% confidence intervals were determined to assess diagnostic value of these biomarkers or the combination of these biomarkers for CSVT diagnosis and chronic progression prediction. The correlation between continuous variables was assessed by the Bravais-Pearson correlation coefficient (in case of normality) or the Spearman’s rank correlation coefficient (in case of non-normality).

All statistical analysis was performed using R Version 4.2.1 (R Foundation for Statistical Computing, Vienna, Austria, URL http://www.R-project.org) and Prism Graph-Pad version 9.01.

The type I error was set to 5% (two-sided). Hence, all inferential results – except for the primary parameter sPECAM-1 for which the sample size estimation was performed, are of descriptive nature only.

## Results

### Baseline characteristics of study patients (Table 1)

**Table.1.** Characteristics, risk factors and laboratory parameters at admission in patients with CSVT and without CSVT

|  | All (n=75) | CSVТ (n=24) | No CSVТ (n=51) | p-value |
| --- | --- | --- | --- | --- |
| Age (mean ± SD, median) | 42.9 ± 17.3 (39) | 44.86 ± 16.1 (43.5) | 40.68 ± 16.5 (39) | 0.25 |
| Female (n,%) | 51 (68) | 18 (78) | 33 (65) | 0.51 |
| BMI (mean ± SD, median) | 23.8 ± 3.7 (22) | 24.3 ± 3.7 (22.5) | 23.3 ± 3.7 (22.0) | 0.46 |
| Immobilization/surgery (n,%) | 5 (6) | 1 (4) | 4 (8) | >0.99 |
| Malignancy (n,%) | 8 (11) | 4 (17) | 4 (8) | 0.67 |
| Thrombophilia (n,%) | 10 (13) | 6 (25) | 4 (8) | 0.24 |
| Previous VTE (n,%) | 3 (4) | 1 (4) | 2 (4) | >0.99 |
| Smokers (n,%) | 30 (40) | 8 (33) | 22 (46) | 0.56 |
| Hormone therapy (n,%) | 22 (29) | 6 (25) | 16 (33) | 0.75 |
| Intracranial hemorrhage (n,%) | 5 (6) | 5 (21) | 0 (0) | **0.05\*** |
| Ischemic stroke (n,%) | 2 (3) | 0 (0) | 2 (4) | 0.49 |
| **Laboratory parameters**<br>(mean ± SD, median) | | | | |
| Creatinin Clearance (mg/dL) | | 0.77 ± 0.24 (0.72) | 0.82 ± 0.3 (0.72) | 0.91 |

Legend Table.1
BMI= Body Mass Index; CSVТ= Cerebral sinus vein thrombosis; VTE= Venous thromboembolism

Seventy-five patients with clinically suspected CSVT were included in the study. Mean age was 42.9 ± 17.3 years (range-19 to 81) and 68% were female. Clinical characteristics, risk factors for CSVT, laboratory parameters and outcome are presented in Table 1. Twenty-four patients (32%) had confirmed CSVT by brain MRI.

Patients with CSVT commonly presented with isolated headache (62.50%), visual impairment (20.83%), seizures and/or focal neurological signs (16.67%). The most common clinical presentation of the control group included headache (78%), dizziness (13.3%) and visual impairment (8%). Neuroimaging revealed thrombus formation in the sinus transversus (71%), sinus sigmoideus (58%), sinus sagittalis superior (37,5%) and with extension to the jugular vein (37.5%). In 67% of patients more than one sinus was occluded and 25% of patients suffered from cortical deep vein thrombosis. Intracranial hemorrhage was present in 21% of patients already on admission as revealed by neuroimaging.

In 51 patients with clinically suspected CSVT, neuroimaging results were negative. Clinical characteristics, risk factors for CSVT and routine laboratory parameters did not differ from patients with proven CSVT (Table 1). Headache was the predominant complaint in patients without CSVT, and none of the patients presented with seizures or focal neurological signs on admission. Interestingly, neuroimaging revealed acute ischemic stroke in two patients without evidence for CSVT and no focal neurological deficits. (silent stroke)

### Thrombus resolution at 3 months and Outcome

Of the 24 patients with acute CSVT, one third (8/24, 33%) had complete resolution of CSVT after 3 months. Further thrombus resolution was observed in additionally 8 patients (33%) by 12 months, leaving one third of all CSVT patients with neuroimaging signs of CSVT beyond 12 months. Other relevant complications included symptomatic epilepsy (N=3/24) and chronic headache(N=6/24). In those patients thrombus persistence was higher than in patients without chronic headache (4/6 versus 2/18, p = 0.01; Odds ratio [OR] 7.0; Relative risk [RR] 3.0 95% CI 0,11 to 0.80)

### Laboratory parameters

#### D-Dimer

D-Dimer-levels were above the cut-off of 0.5mg/L in 35/75 patients (46%) including 16 patients without CSVT. In 4 patients with CSVT D-Dimer levels were not elevated (16%), as in patients with ischemic stroke (N=2). Mean baseline D-Dimer-levels were significantly higher in the CSVT group compared to the control group without CSVT (p-value <0.001; Table.2). D-Dimer levels > 0.5mg/L had a diagnostic sensitivity of 83% and specificity of 73% for CSVT. D-Dimer levels did correlate with sPECAM-1 baseline levels [r=0.50 (95% CI 0.24-0.70); p-value <0.001] but not with creatinine-clearance, sICAM-1 and sVCAM-1 levels and the number of affected sinuses.

**Table. 2.** Laboratory biomarkers at admission in patients with CSVT and without CSVT

| **Laboratory biomarkers**<br>(mean ± SD, median) | **All (n=75)** | **CSV T (n=24)** | **No CSV T (n=51)** | **p-value** |
| --- | --- | --- | --- | --- |
| D-Dimer mg/L | 1.12 ± 2.02 (0.5) | 2.468 ± 3.29 (1.67) | 0.519 ± 0.28 (0.45) | **<0.001\*** |
| sPECAM-1 ng/mL | 167.3 ± 65.1 (149.3) | 246.8 ± 55.6 (225.7) | 137.2 ± 36.1 (137.5) | **<0.001\*** |
| sICAM-1 ng/mL | 823.7 ± 133.8 (811.4) | 871 ± 244.7 (883.2) | 775.1 ± 369.1 (826.4) | 0.09 |
| sVCAM-1 ng/mL | 818.3 ± 277.1 (859.1) | 903.5 ± 137 (887.5) | 827.9 ± 123.0 (818.6) | 0.25 |
| **Laboratory biomarkers at 3 months after index event in patients with CSV T**<br>(mean ± SD, median) | | | | |
| D-Dimer mg/L | 0.62 ± 0.8 (0.35) | | | |
| sPECAM-1 ng/mL | 116.8 ± 32.7 (121.7) | | | |
| sICAM-1 ng/mL | 843 ± 96.7 (871.1) | | | |
| sVCAM-1 ng/mL | 779.4 ± 217.7 (830.5) | | | |

Legend Table.2
CSV T= Cerebral sinus vein thrombosis; sICAM-1= soluble intercellular adhesion molecule-1; sPECAM-1= soluble platelet endothelial cell adhesion molecule-1; sVCAM-1= soluble vascular adhesion molecule-1.

#### sPECAM-1

Mean baseline sPECAM-1 levels were significantly higher in patients with CSVT compared to patients without CSVT. (p-value <0.001; Table 2) Elevated sPECAM-1 levels above 198.7ng/mL had a diagnostic sensitivity of 95.7% and a specificity of 90.9% for detecting CSVT. (FPR: N=3, 4%; FNR: N=2, 9%).

Baseline sPECAM-1 levels correlated with D-Dimer (see above) and sICAM-1 levels [r = 0.44 (95% CI 0.16 to 0.66); p-value = 0.002]. There was no difference in the number of affected cerebral sinus veins or in isolated cortical vein and sinus vein thrombosis and receptor protein levels.

#### sICAM-1

There was no difference in sICAM-1 levels between patients with or without CSVT (p-value = 0.09; Table 2). sICAM-1 levels only correlated with sPECAM-1 at baseline (p=0.02). There was no difference in the number of affected cerebral sinus veins or in isolated cortical vein and sinus vein thrombosis.

Baseline sICAM-1 levels were significantly higher in patients with persistent neuroimaging signs of cerebral sinus vein thrombosis beyond 12 months (846 vs 1069 ng/ml; p-value <0.001)

#### sVCAM-1

There was no difference in sVCAM-1 levels between patients with or without CSVT (p = 0.25; Table.2). sVCAM-1 levels didn’t correlate with any other parameter at baseline and follow up.

### D-Dimer, sPECAM-1, sICAM-1 and sVCAM-1 in predicting CSVT (Table 3)

**Table. 3.** Values from ROC Curve Analyses.

|  | Data by Group Comparison |  |
| --- | --- | --- |
|  | CSVT vs Control - BL | Resolution vs. Chronic |
| **D-Dimer** |  |  |
| -AUC (95% CI) | 0.85 (0.62-0.93) | 0.73 (0.51-0.98) |
| -Sensitivity | 50.00 | 66.67 |
| -Specifity | 93.75 | 62.50 |
| -PPV | 80.00 | 76.92 |
| -NPV | 78.95 | 50.00 |
| -Likelihood ratio | 8.00 | 3.750 |
| -p-value | **<0.001\*** | 0.08 |
| **sPECAM-1** |  |  |
| -AUC (95% CI) | 0.95 (0.90- 1.00) | 0.99 (0.86- 1.00) |
| -Sensitivity | 95.65 | 100.00 |
| -Specifity | 90.91 | 86.67 |
| -PPV | 94.12 | 80.00 |
| -NPV | 87.50 | 100.00 |
| -Likelihood ratio | 10.52 | 15.00 |
| -p-value | **<0.001\*** | **<0.001\*** |
| **sVCAM-1** |  |  |
| -AUC (95% CI) | 0.60 (0.43- 0.77) | 0.66 (0.41-0.92) |
| -Sensitivity | 47.83 | 62.5 |
| -Specifity | 72.73 | 79.10 |
| -PPV | 42.86 | 26.32 |
| -NPV | 74.47 | 94.64 |
| -Likelihood ratio | 1.754 | 4.688 |
| -p-value | 0.25 | 0.2 |
| **sICAM-1** |  |  |
| -AUC (95% CI) | 0.65 (0.49- 0.81) | 0.94 (0.85- 1.00) |
| -Sensitivity | 43.48 | 75.00 |
| -Specifity | 81.82 | 93.33 |
| -PPV | 55.56 | 64.55 |
| -NPV | 75.44 | 95.31 |
| -Likelihood ratio | 2.391 | 11.25 |
| -p-value | 0.08 | **<0.001\*** |
| **D-Dimer+sPECAM-1** |  |  |
| -AUC (95% CI) | 0.99 (0.96-1.00) | 0.99 (0.96-1.00) |
| -Sensitivity | 100.00 | 100.00 |
| -Specifity | 96.08 | 96.00 |
| -PPV | 92.31 | 72.73 |
| -NPV | 100.00 | 100.00 |
| -Likelihood ratio | 25.5 | 25.5 |
| -p-value | **<0.001\*** | **<0.001\*** |

Legend Table.3
AUC= Area under the curve; BL= Baseline; CI= Confidence interval; CSVT= Cerebral sinus vein thrombosis; NPV= Negative predictive value; PPV= Positive predictive value; ROC= Receiver Operating Characteristics; sICAM-1= soluble intercellular adhesion molecule-1; sPECAM-1= soluble platelet endothelial cell adhesion molecule- 1; sVCAM-1= soluble vascular adhesion molecule-1

Using ROC analyses, sPECAM-1 and D-Dimer had the highest AUC (0.89 and 0.86, respectively) in identifying patients with CSVT (Fig. 2, Table 3) Logistic regression analysis revealed that the combination of D-Dimer and sPECAM-1 yielded the best AUC (0.994; 95% CI 0.98 to 1.000; p □< □0.001) with a negative predictive value (NPV) of 95.65% and a positive predictive value (PPV) of 95.45% (Table 3) The combination of D-Dimer (cut-point □> □0.5 mg/L) and sPECAM-1 (cut-off> 198.7ng/mL) resulted in a reduction of patients with “false-negative” D-Dimer levels from 4 to 0 cases.

**Fig.2.**
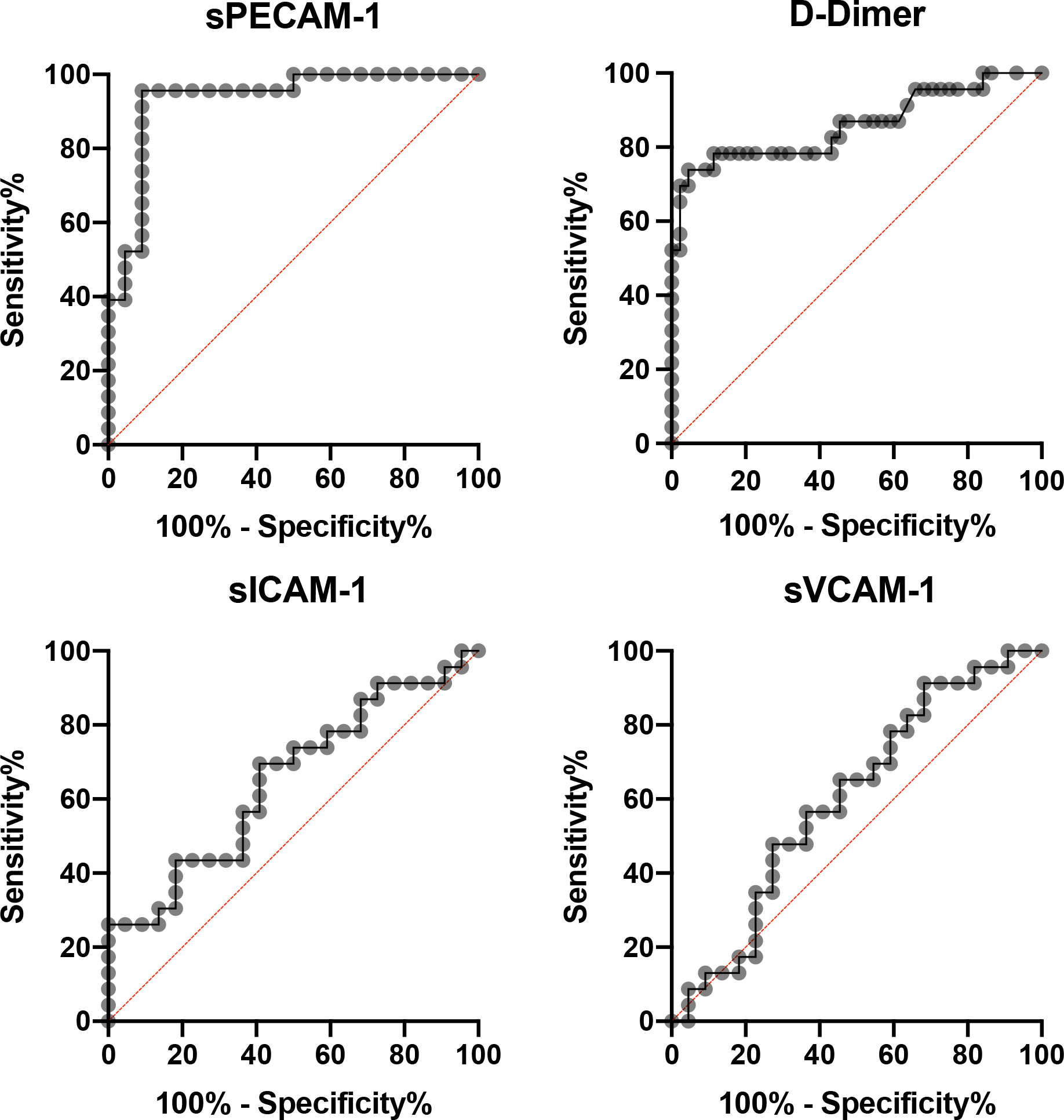
Receiver operating curves for predicting CSVT at BL

In patients with isolated headache and without focal neurological signs and symptoms (n = 15) sPECAM-1 > 198.7ng/mL had an AUC of 0.94 (95% CI 0.86 - 1.000)). Again, the combination of sPECAM-1 and D-Dimer increased the AUC to 0.99 (95% CI 0.97-1.000).

### D-Dimer, sPECAM-1, sICAM-1 and sVCAM-1 in predicting delayed thrombus resolution (Table 3)

ROC curves in predicting delayed thrombus resolution and chronification are shown in Fig. 3. and Fig. 4. AUC was 0.61, 0.83 and 0.80 for D-Dimer, sPECAM-1 and sICAM-1, respectively.

**Fig.3.**
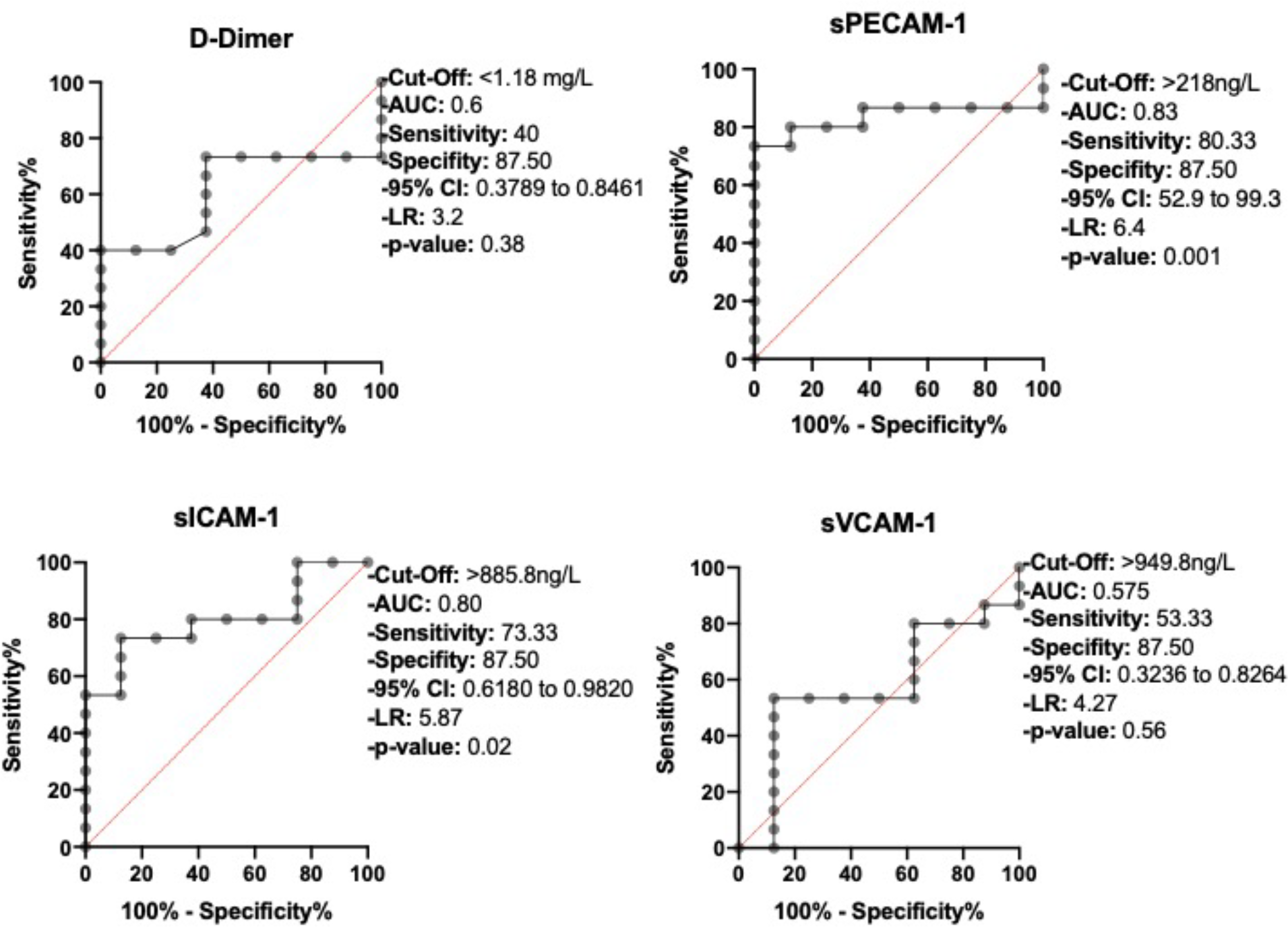
Receiver operating curves for predicting delayed thrombus dissolution

**Fig.4.**
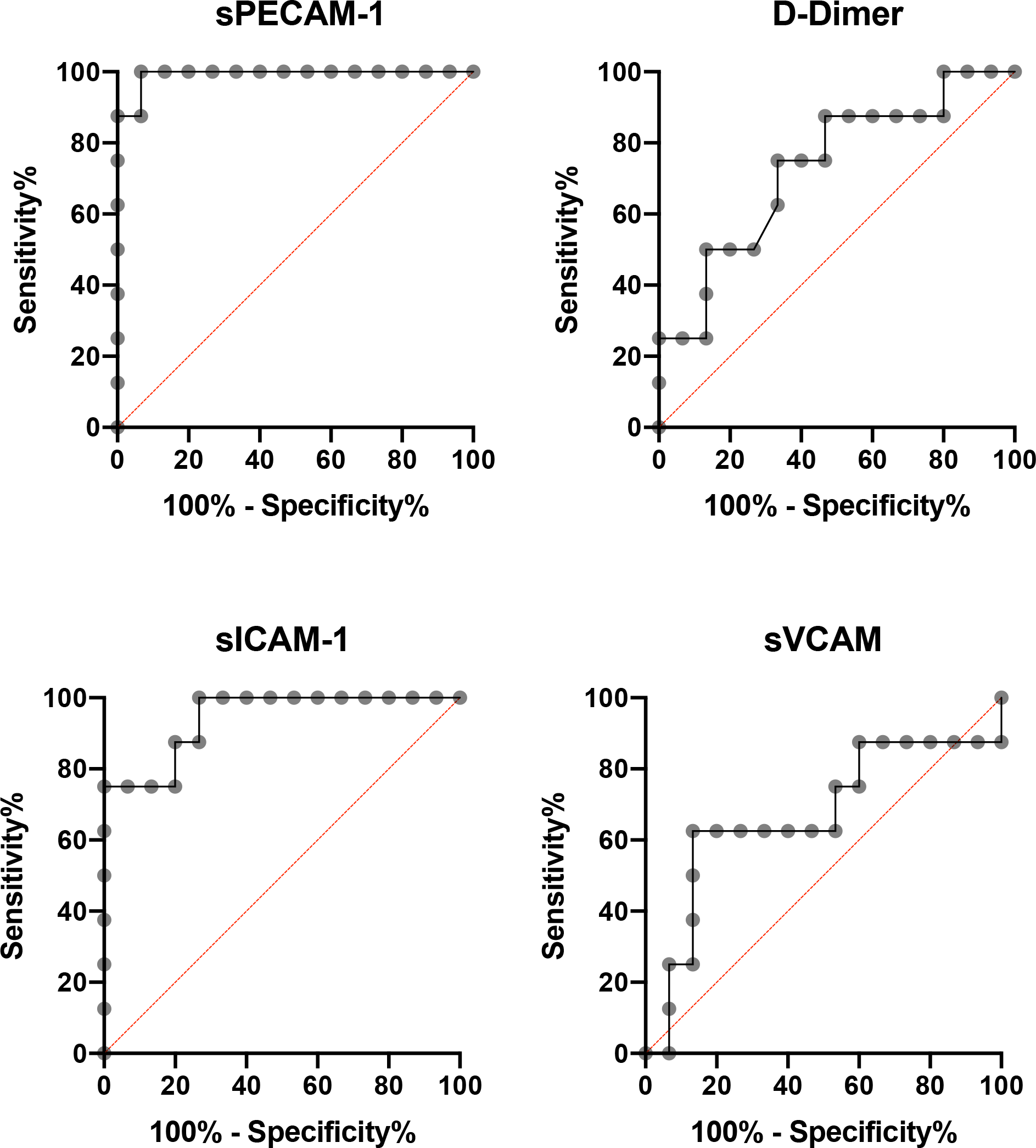
Receiver operating curves for predicting chronification in CSVT

## Discussion

In this study we found that (1) baseline sPECAM-1 levels were significantly higher in patients with CSVT compared to patients without CSVT, and (2) sPECAM-1 may qualify as a reliable biomarker not only for predicting CSVT, but also for predicting delayed radiologic thrombus resolution. To our knowledge this is the first study to investigate the role of endothelial receptor proteins in CSVT. Interestingly, the combination of D-Dimer and sPECAM-1 plasma levels was better in predicting chronic CSVT compared to each biomarker alone, especially D-Dimer.

D-Dimer is the only routinely used diagnostic biomarker in patients with suspected VTE, ^12,24^ and PE or DVT can be safely ruled out in low risk situations (Wells score <4) without the need for further imaging.^9,25,26^ In contrast, data about the diagnostic accuracy of D-Dimer in patients with CSVT remain controversial. Two systematic reviews showed that patients with clinical isolated headaches or symptom duration > 7 days have often negative D-Dimer,^9,11^ In our study D-Dimer (Cut-off 0.5mg/L) had a sensitivity of 82.61% and a specificity of 72.73%, which confirms that isolated D-Dimer levels are not useful as a diagnostic approach. Interestingly, D-Dimer levels >0.5 had a sensitivity of 100% and a specificity of 52% in patients with visual (diplopia or anopia) or other focal neurological disorders (in our case focal paresis or hemiparesis), which is in line with previous studies (mean sensitivity between, 93% and 97.8% respectively). ^9,11^ The high rate of false positive results of D-dimer may be explained by elevated levels observed in advanced age, during pregnancy, smoking, heart disease or acute inflammation.^27,28^ Therefore D-Dimer measurement is not recommended in the guidelines for the diagnosis of cerebral sinus vein thrombosis to avoid unnecessary imaging. D-Dimer is often influenced by pro-inflammatory condition in contrast to sPECAM-1 ^13,19^ Therefore, we think additional sPECAM-1 measurement in patients with suspected CSVT can be helpful especially in patients with isolated headache or patients with unspecific inflammatory conditions (pregnancy or neoplastic disease) due to its robustness against pro-inflammatory cytokines. Another finding in our study is that sPECAM-1 and sICAM-1 were associated with thrombus persistence Negative parameters of sPECAM-1 could contribute to save costs and avoid unnecessary imaging.

sPECAM-1 and sICAM-1 have been previously described to predict chronic VTE.^13,15,16^ Though its role in CSVT is still unclear due to the unique anatomical particularities of the dural sinus. The dural sinus veins contain no muscular tissue, possess no valves and the walls are composed of dura mater lined by endothelium^29,30^, which could explain the high rate of chronification in CSVT. In our study sPECAM-1 and sICAM-1 were associated with thrombus persistence.

Further neuropathological examinations could help identify the role of sPECAM-1 and sICAM-1 in thrombus resolution at the side of sinus veins thrombosis. We therefore retrospectively stained 7 thrombi of patients who died from sinus vein thrombosis. The staining pattern revealed that PECAM-1 correlated significantly with granulocytes and monocytes in the thrombus which confirmed that PECAM-1 plays an important role in thrombus resolution not only in deep vein thrombosis but also in cerebral sinus vein thrombosis despite the anatomical differences of the endothelial wall. (unpublished data)

Our patients had anticoagulant treatment for at least 3 months. Although high rates of recanalization was detected in the first 3 months after the index event in large and small cohorts of patients with CSVT, about two third in our cohort had not resolved the thrombus after 3 months.^31,32^ Recent guidelines do not give a clear recommendation for the ideal duration of oral anticoagulation, which often limits young patients in daily activities and sports. Longitudinal studies about monitoring the thrombus development and resolution of CSVT are rare.^31^ We found that sPECAM-1 is able to identify patients with risk of delayed thrombus and may be beneficial and helpful to individually decide the ideal duration of oral anticoagulation of the patient and reduce the rate of chronic sinus vein thrombosis.

The present study has important limitations. First, the study is based on a relatively small sample size. But sample size calculation based on data in Kellermair et al. ^13^ showed a sufficient power of 90% at a type I error of 5%. Secondly, we didn’t include pregnant women, patients with postpartal CSVT and patients with septic sinus vein thrombosis, which limits generalizability of our results. Thirdly, although there is evidence that lysis of the thrombus and recanalization of venous segments are typically observed in the first weeks, ^33^ the definition of thrombus persistence or delayed thrombus resolution is arbitrarily chosen. The definition of “chronic sinus thrombosis” is also not clearly standardized and chosen similar to the definition of chronic deep vein thrombosis (thrombotic residues after 6-12 months)^34,35^ as still significant thrombotic residues after 12 months in neuroimaging. Fourth, we did not include a control group of patients without headache.

In conclusion sPECAM-1, sICAM-1 and D-Dimer can be helpful parameters for the acute diagnosis of CSVT and can predict delayed thrombus resolution and therefore, could help identify the ideal duration of anticoagulation and avoid excessive imaging checks.

## Supporting information

Checklist

## Data Availability

The data that support the findings of this study are available on request from the author

## Authors’ Roles

1. Research Project: A Conception, B. Organization C Execution
2. Statistical Analysis: A Design, B Execution C Review and Critique
3. Manuscript: A Writing of the first draft B Review and Critique

**Lukas Kellermair:** 1A, 1B, 1C, 2A, 2B, 3A; **Christoph Höfer:** 1C, 2C, 3B; **Matthias W.G. Zeller:** 1C, 2C, 3B; **Christa Kubasta:** 1B, 2C, 3B; **Bandke, Dave:** 2C, 3B; **Serge Weis**: 1B, 2C, 3B; **Jörg Kellermair**: 2C, 3B; **Thomas Forstner:** 2A, 2B, 3B; **Raimund Helbok**: 2C, 3B; **Milan R. Vosko:** 1A, 1C, 2C, 3B

